# Inferred resolution through herd immmunity of first COVID-19 wave in Manaus, Brazilian Amazon

**DOI:** 10.1101/2020.09.25.20201939

**Authors:** Thomas A. A. Prowse, T. Purcell, Djane C. Baía-da-Silva, V. Sampaio, Wuelton M. Monteiro, James Wood, I. Mueller, Jodie McVernon, Marcus V. G. Lacerda, Joshua V. Ross

**Affiliations:** School of Mathematical Sciences, The University of Adelaide, Adelaide, South Australia 5005, Australia; Victorian Infectious Diseases Reference Laboratory Epidemiology Unit at The Peter Doherty Institute for Infection and Immunity, The University of Melbourne and Royal Melbourne Hospital, Melbourne, Victoria 3000, Australia; Fundação de Medicina Tropical Dr Heitor Vieira Dourado, Manaus, Brazil; Fundação de Vigilância em Saúde; Fundação de Medicina Tropical Dr Heitor Vieira Dourado, Manaus, Brazil; Universidade do Estado do Amazonas; Fundação de Medicina Tropical Dr Heitor Vieira Dourado, Manaus, Brazil; School of Population Health, UNSW Sydney, Australia; Population Health and Immunity Division, Walter + Eliza Hall Institute, Parkville, Victoria, Australia; Department of Medical Biology, FMDHS, University of Melbourne, Parkville; Department of Parasites and Insect Vectors, Institut Pasteur, Paris, France; Centre for Epidemiology and Biostatistics, Melbourne School of Population and Global Health, The University of Melbourne; Infection and Immunity Theme, Murdoch Childrens Research Institute, Parkville; Instituto Leônidas & Maria Deane, Fiocruz; Fundação de Medicina Tropical Dr Heitor Vieira Dourado, Manaus, Brazil

**Author notes:** Joshua V. Ross, School of Mathematical Sciences, The University of Adelaide, Adelaide, South Australia 5005, Australia. equal first authors (Thomas A. A. Prowse, T. Purcell). equal last authors.

## Abstract

**Background:** As in many other settings, peak excess mortality preceded the officially reported ‘first wave’ peak of the COVID-19 epidemic in Manaus, Brazil, reflecting delayed case recognition and limited initial access to diagnostic testing.

**Methods and Findings:** To avoid early information bias, we used detailed age and gender stratified death certificate and hospitalisation data to evaluate the epidemic’s trajectory and infer the cause of its decline using a stochastic model. Our results are consistent with heterogenous transmission reducing from mid-April 2020 due to the development of herd immunity. Relative to a baseline model that assumed homogenous mixing across Manaus, a model that permitted a self-isolated population fraction reduced the population-wide attack rate required to drop the effective reproduction number below one from 62 % to 47 %, and reduced the final attack rate from 86% to 65%. In the latter scenario, a substantial proportion of vulnerable, older individuals remained susceptible to infection.

**Conclusions:** Our models indicate that the development of herd immunity amongst the mixing proportion of the Manaus population had effectively halted the COVID-19 epidemic by late July 2020. Given uncertainties regarding the distancing behaviours of population subgroups with different social and economic characteristics, and the duration of sterilising or transmission-modifying immunity in exposed individuals, we conclude that the potential for epidemic outbreaks remains, but that future waves of infection are likely to be much less pronounced than that already experienced.

## Introduction

Globally, marked differences have been observed in morbidity and mortality due to SARS-CoV-2 infection. This variability mostly reflects the extent and timeliness of spontaneous and imposed changes in social mixing and the sensitivity of surveillance systems to detect cases and deaths [1]. Many countries that successfully constrained the initial epidemic through distancing measures are now experiencing second waves in still-susceptible populations [2].

Estimates of *R*_0_ for SARS-CoV-2 in the range 2 to 6 [3, 4] suggest that population immunity of approximately 50-80% is required to achieve herd protection. However, heterogeneous behaviour, infectivity and immunity within subpopulations could plausibly decrease this threshold to 10-20% [5]. Controversy remains regarding the extent of population exposure required to achieve such constraint [6].

Comparison of population attack rates to inform this question is made challenging by imperfect case ascertainment, compounded by limited diagnostics and overwhelmed health systems, particularly in high incidence settings. Assessment of epidemic activity therefore requires the use of less biased metrics than confirmed case reports. Excess mortality is an objective measure which, with cause-of-death certification, can be used as an indicator of direct and indirect COVID-19 associated mortality [7]. With hospitalisations data, it can inform retrospective estimation of cumulative cases and deaths [8].

Brazil experienced a severe first wave of COVID-19 disease, with mass mortality reported in many states, mainly in the north where seasonality of respiratory infections contributed to higher vulnerability. A socialized health system provided free and global access to tertiary care hospitals, but inequalities might explain different mortality rates in the population [9]. In Manaus, the highly urbanised capital of Amazonas state, the first case of COVID-19 was reported on 13 March 2020 [10]. By 11 August 2020, 37,597 cases and 2,051 deaths were reported [11].

However, burial and death records indicate far higher mortality than official reports, suggesting late recognition of importation and underreporting. Previous studies have assumed that the first wave in Manaus was significantly mitigated by non-pharmaceutical interventions (NPIs) [12]. While these restrictions may have partly constrained early transmission, local reports indicate that implementation was highly variable [13]. Moreover, a possible role for immunity is suggested by the observation of declining cases and deaths over a period in which restrictions were officially eased.

We use death certificate and hospitalisation records to parameterise an epidemiological model of the COVID-19 epidemic in Manaus. The model allows inference of age- and gender-stratified infection-fatality ratios to explore evidence for development of herd immunity as a driver of local epidemic resolution, with implications for the ongoing risk posed by SARS-CoV-2 to this population.

## Methods

### Excess mortality and reported causes

Death certificate data from January 2015 to July 2020 were sourced from the Brazilian Ministry of Health Mortality Information System (SIM). Information on age, gender, and cause and date-of-death was recorded. Cause of death was reported using the World Health Organization’s (WHO) International Classification of Diseases 10^th^ revision (ICD-10). The 2010 population census and the 2019 municipal population estimates were accessed from the Brazilian Institute of Geography and Statistics (IBGE). This project was approved by the *Fundação de Medicina Tropical Dr Heitor Vieira Dourado* Ethics Review Board (Approval 4.033.218).

Background mortality was calculated by averaging the number of deaths observed per week, commencing on January 1 each year, for the previous 5 years (2015-2019). Background mortality was further stratified by gender and into 5-year age bands, culminating at 75+. For each subgroup, we subtracted this five-year average from deaths observed for the period between March 19 and June 24 of 2020 to determine excess deaths. This period was selected as excess mortality was first observed in the week beginning on March 19 and the data returned to baseline background mortality in the week beginning on June 25.

The age and gender structure reported in the 2010 population census was used to estimate the 2019 population size for each age and gender class. Population estimates were aggregated into 5-year age bands. The 5-year population estimates for 2019 were used to determine excess mortality, as a proportion of the population.

The ICD-10 codes assigned to each death certificate were aggregated into 7 categories: diseases of the respiratory system (J00-J99), circulatory system (I00-I99), endocrine system (E00-E99), unattended (R98) and unknown cause of death (R99), coronavirus infection (B34.2) and all remaining codes were categorised as ‘other’. All analyses were performed using R software.

### Hospitalisation data

Hospitalisation data for patients admitted to all hospitals (private and public) in Manaus from January 2020 to July 2020 was accessed from the Influenza Surveillance Information System (SIVEP-Gripe). Data entry in this system is compulsory by law. A deidentified line list included patient demographics, comorbidities, clinical symptoms, investigations, clinical management and outcome data.

Daily admissions to both hospital and intensive care units (ICU) were aggregated each week in 2020 and stratified into 3 age bands (0-29, 30-64 and 65+). For each week, the proportion of hospital and ICU admissions within each age group was determined. Between April and July, the proportional distribution of hospital and ICU admissions across the three categories remained fairly stable, with no evidence of rationing of service access on the basis of age.

### Epidemiological model

We developed a stochastic, discrete-time, susceptible-infected-recovered epidemiological model for Manaus which we calibrated using daily time-series data from the death-certificate and hospitalisation records. The model assumed an initial population size equal to that of Manaus in 2019 (2,182,761), which was split into *N*=36 age/gender groups based on proportions reported in the 2010 census (i.e., 5-year age classes up to an age of 75 years, and then a pooled age class for all individuals aged 75 years and over). To allow inference on the background mortality rate in each age/gender group, the model was initiated on January 1, 2020, approximately two months prior to the introduction of COVID-19. The model was terminated after *n=*202 days on July 20, 2020, to ensure complete reporting of both death certificates and hospitalisations over the modelled period. The model detailed below was fitted within a Bayesian framework using *JAGS* (v. 4.3.0) software [14] and a mixture of informative and uninformative priors (for full details of the model code, including details of all prior distributions, see Supplementary Appendix S1).

### COVID-19 introduction and virus transmission

We initialised the model by allowing importation of COVID-19 cases into Manaus over the week beginning March 5, 2020 (one week before the first reported case), and inferred an importation model such that the number of introduced cases into each group in each day of this first week arose from a Poisson process with a common mean inferred from the data. To model community transmission, we inferred a reproduction number on day *t* that was modified by an index of human mobility

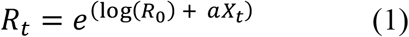

where *R*_0_ is the basic reproduction number, *X*_*t*_ is the mobility covariate and *a* is the inferred coefficient. We derived the time-series *X*_*t*_ by first averaging daily data for five separate indices of community mobility (available as the mobility change relative to baseline for retail/recreation, grocery/pharmacy, parks, transit stations and workplaces, accessed from www.google.com/covid19/mobility), and then smoothed the series with a 7-day moving-average smoother. We then assumed the effective reproduction number 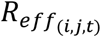 in group *i* due to mixing with group *j* on day *t* is

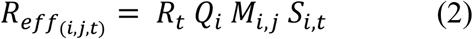

where *Q*_*i*_ is the susceptibility of group *i* to infection, *M*_*i,j*_ is the rate of mixing between the two groups, and *S*_*i,t*_ is the time-varying proportion of susceptible (previously uninfected) individuals remaining in the focal group. Based on previous studies [15, 16], we used prior means for the susceptibilities of 0-15, 15-60, and 60+ year old age classes of 0.5, 1.0, and 1.3, respectively.

Mixing between groups was governed by an age-structured mixing matrix reported previously for Brazil [17], which we corrected to be symmetrical (by averaging the upper and lower triangles), scaled to a mean of 1/*N*, and applied equally to both genders. The expected number of new cases in each group each day was then calculated as

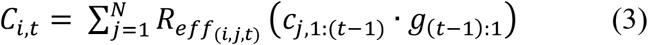

where *c*_*j*,1:(*t*−1)_ is the case-history vector for group *j* and *g*_(*t*−1):1_ is the portion of the generation interval distribution relevant to those cases. We used the same generation interval distribution as Mellan et al. [12] which concentrated >99% of an individual’s infectivity within the first 3 weeks of infection (median generation interval=5 days).

To account for heterogeneity in transmission, we assumed the offspring distribution in group *i* due to mixing with infectious individuals from group *j* on day *t* was governed by a negative binomial distribution with mean equal to *C*_*i,t*_ and variance equal to *C*_*i,t*_(1 + *C*_*i,t*_/∅_*i,t*_), where

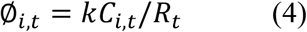

and *k* is the overdispersion parameter, such that smaller values of *k* represent greater transmission heterogeneity (i.e., the more transmission is due to a small number of people, including by so-called “superspreaders”). Given no previous study has documented strong evidence of different susceptibility between genders, we first generated the expected number of new cases in each age class (regardless of gender), and then assumed gender-specific cases arose from a binomial distribution with probability equal to the proportion of the total susceptible individuals for that age class attributable to each gender.

### Background and COVID-induced mortality

We developed a model for the expected number of deaths per day which was comprised of three components. First, we modelled the background (pre-COVID-19) death rate *d*_*i*_ estimated separately for each group. Second, available data on the time from symptom onset to death for confirmed COVID-19 cases (from the Manaus hospitalisation records) were used to infer a 4-parameter (*y*_0_, *u*_min_, *u*_0_, *b*) mortality distribution (*m*) as a function of the time *x* since symptom onset (*x* ∈ {-5, …, *N*-5}) of the form

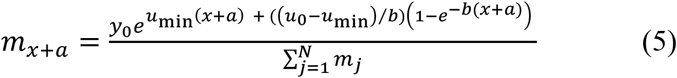

where *a* is the average time from infection to symptoms, which was fixed at 5 days [3, 18, 19]. Finally, we modelled the infection-fatality ratio in each group *i* (*IFR*_*i*_) as a log-linear function of age and gender, with prior distributions on this components’ parameters based on a recent meta-analysis of COVID-19-induced mortality [20]. Together, these components yielded the following model for the expected number of deaths

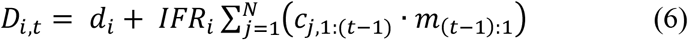

and the observed number of deaths each day was assumed to arise from a Poisson distribution with mean equal to *D*_*i,t*_.

### Hospitalisations due to COVID-19

Hospitalisations were modelled in a similar way to COVID-induced deaths, in that available data on the time from symptom onset to hospitalisation were used to infer a 4-parameter hospitalisation distribution (*h*) of the same functional form as that used for the mortality distribution above. We modelled the infection-hospitalisation ratio in each group *i* (*IHR*_*i*_) as a logistic function of age and gender. Daily hospitalisations were assumed to arise from a Poisson distribution with mean equal to

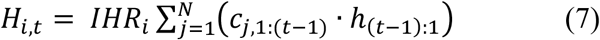

### Model scenarios

Given no data were available on whether some portion of the Manaus population has been self-isolating since March 2020, we initially fitted a ‘baseline’ model to death-certificate and hospitalisation data which assumed homogeneous age-structured mixing across the entire Manaus population. We compared these model outcomes to those from a second model that permitted a self-quarantined proportion (*P*) of the population which could not be exposed to the SARS-CoV-2 virus. Communication with local experts including authors on this paper suggested that more wealthy residents of Manaus had been able to greatly reduce social interactions during the first epidemic wave and avoid exposure to the virus. We reviewed detailed socio-demographic data on Manaus [21], but were not able to source quantitative estimates of the relevant population fraction. However, we considered that this would not exceed the upper 2 income quintiles (40%) of the population but was likely to be greater than the wealthiest 10%, and accordingly, chose a non-informative uniform (0.1,0.4) prior on this population fraction.

### Herd-immunity threshold

To estimate the herd-immunity threshold for the baseline model scenario, we calculated the discrete-time next-generation matrix from the mixing matrix and posterior means for *R*_0_ and age-structured susceptibilities *Q*. We then calculated the deterministic reproduction number (*R*) as the dominant eigenvalue of this matrix, and estimated the herd-immunity threshold as 1 − 1/*R*. For each model, we also derived another estimate of this threshold that incorporated the inferred age/sex distribution of cases, by calculating the population-wide attack rate at which the expected number of offspring per case (*R*_*eff*_ multiplied by the normalised case-infectivity vector) fell below one.

## Results

We calculated 3,457 excess deaths in Manaus, Brazil, between 19 March and 24 June 2020 (Supplementary Table 1) representing 0.16% of the city’s population. Males 30 years and over experienced greater excess mortality than females; individuals aged 75 years or more accounted for 39% of the excess (Fig 1A). During this period, 7% of the 75+ male population in Manaus died (Fig 1B). COVID-19 deaths were first reported from 26 March and increased weekly thereafter in keeping with improved access to diagnostics and/or increasing prevalence (Fig 2), comprising 53% of the total excess. Other reported causes of death included respiratory diseases, unattended and unknown causes of mortality, cardiovascular, endocrine and cancer-related mortality, the majority of which were compatible [22] with a clinical diagnosis of COVID-19 or are known comorbidities associated with severe outcomes (Fig 2).

**Fig 1.**
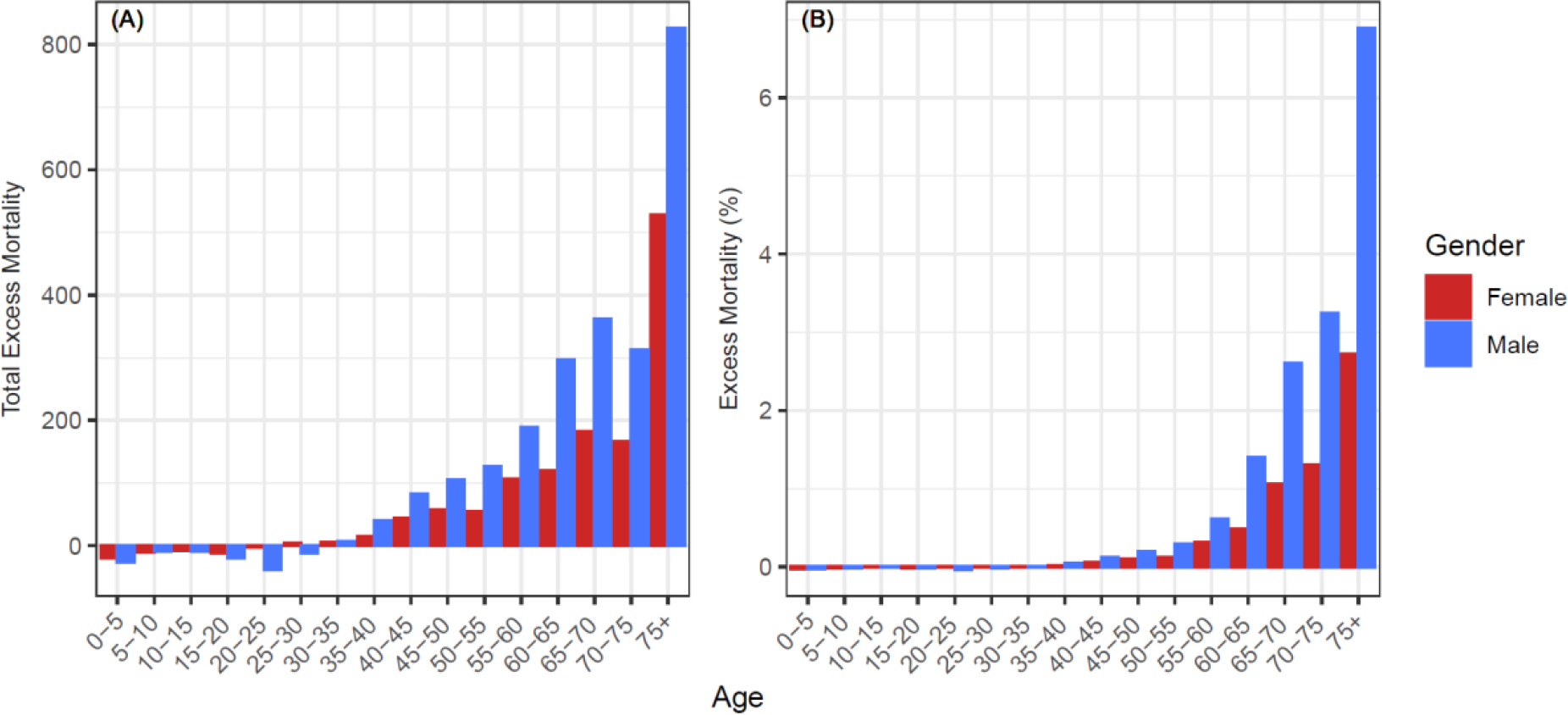
Excess mortality in Manaus during the COVID-19 epidemic. (a) Total excess mortality by age and gender for the period between March 19 and June 24 of 2020. Excess mortality is the 2020 weekly observed mortality less the expected mortality summed for the period between March 19 and June 24 of 2020 for each subgroup. (b) Excess mortality for the period between March 19 and June 24 of 2020 divided by the estimated 2019 subgroup populations to obtain population proportions.

**Fig 2.**
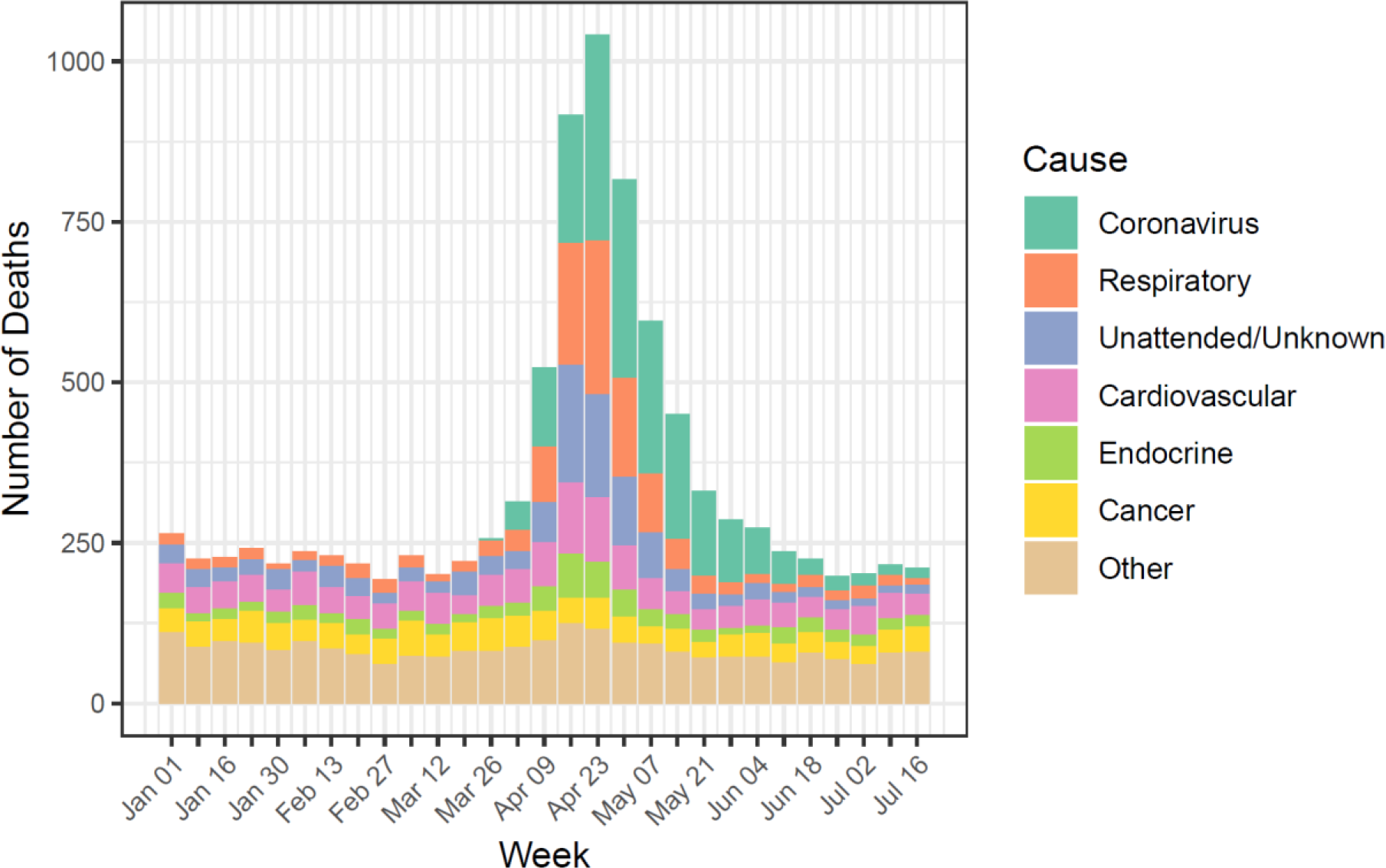
Weekly mortality in Manaus by reported cause of death during the COVID-19 epidemic. Observed weekly mortality for the period between January 01 and July 22 of 2020 aggregated by major ICD-10 categories of attributed causes of death.

Stochastic transmission models captured the observed peak in excess deaths in late April, including age and gender variation (Fig 3, Figs S1 and S2). The models also captured synchronous peaks in hospitalisations (Fig 3, Figs S3 and S4), although model fit to these data was poorer, potentially reflecting delayed recognition of COVID-19 cases and/or capacity exceedance during the epidemic peak.

**Fig 3.**
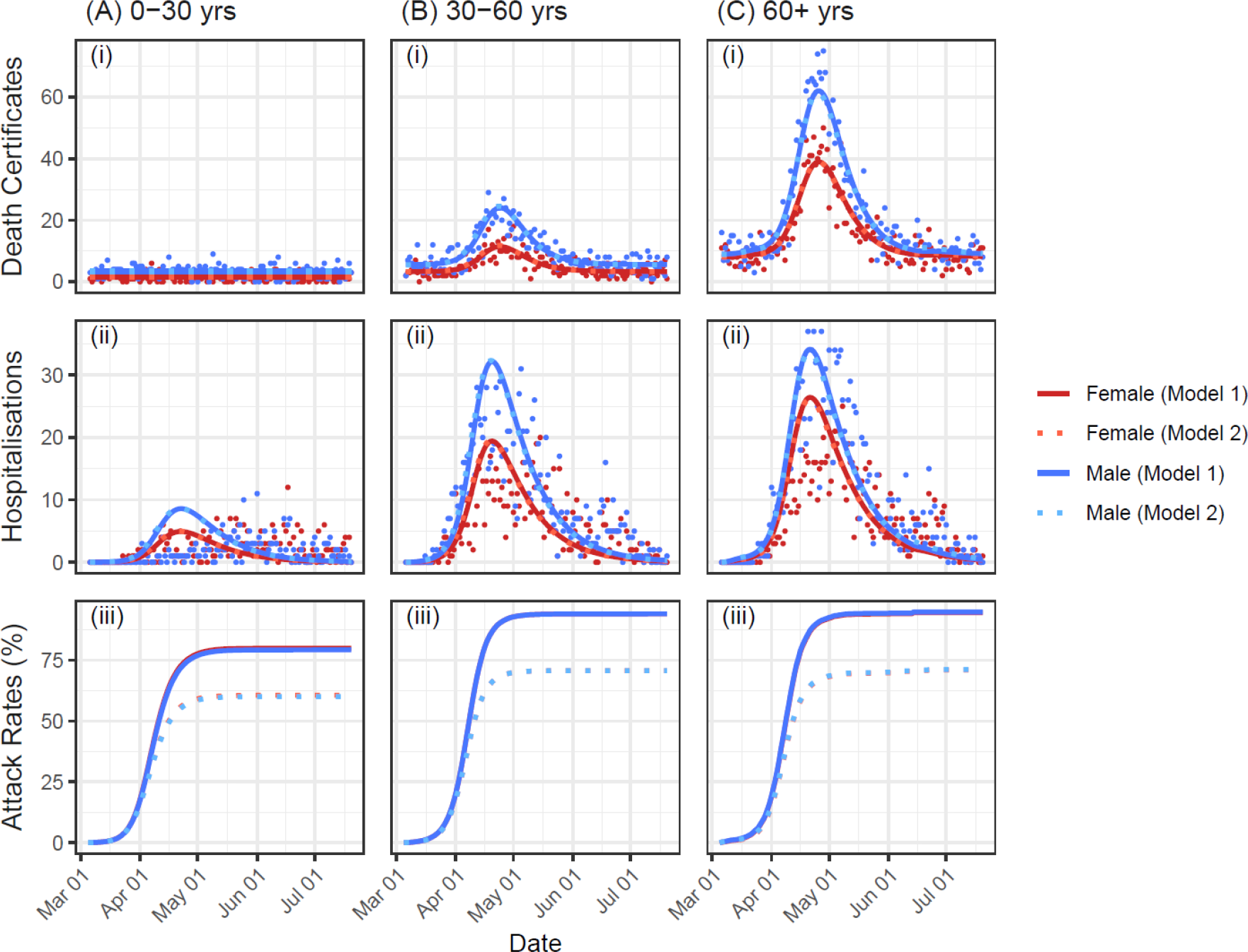
Integrated model fits by gender and model. Results are pooled at the level of three aggregated age classes: (A) 0-30 years; (B) 30-60 years; and (C) 60+ years. For each age class, panels (i) and (ii) show the fit of the model (lines) to the reported number of deaths and hospitalisations per day (points), while panel (iii) shows inferred attack rates. Note that lines showing the model fits overlap in panels (i) and (ii), and attack rates for males and females overlap in panel (iii).

Despite their different assumptions about the proportion of the Manaus population at risk, the two model scenarios yielded equivalent fits to the data (Fig 3A, B). The baseline model estimated heterogenous transmission (k=0.065 [0.047, 0.093], mean [95 % credible intervals]), a mean time-to-death of 13 days (Fig S5), and inferred a population-wide attack rate of 85.7 [84.6, 86.7]% by 20 July. Attack rates were lowest in younger age classes (Fig 3A,i), reflecting lower susceptibility assumed for people under 15 years.

Infection fatality rate (IFR) estimates for the baseline model ranged from almost zero for both genders aged 30-35 years, to 3.0 [2.8, 3.3]% and 7.4 [6.8, 8.0]% for females and males over 75 years, respectively (Fig 4). At a population level, reduced mobility resulted in variable *R*_*t*,_ while the expected offspring per case fell below one after April 12 (Fig S6), when the estimated attack rate was 61.8 [61.1, 62.6]%. The traditional estimate of the herd-immunity threshold based on the discrete-time next-generation matrix calculated from this model was 71.8 %.

**Fig 4.**
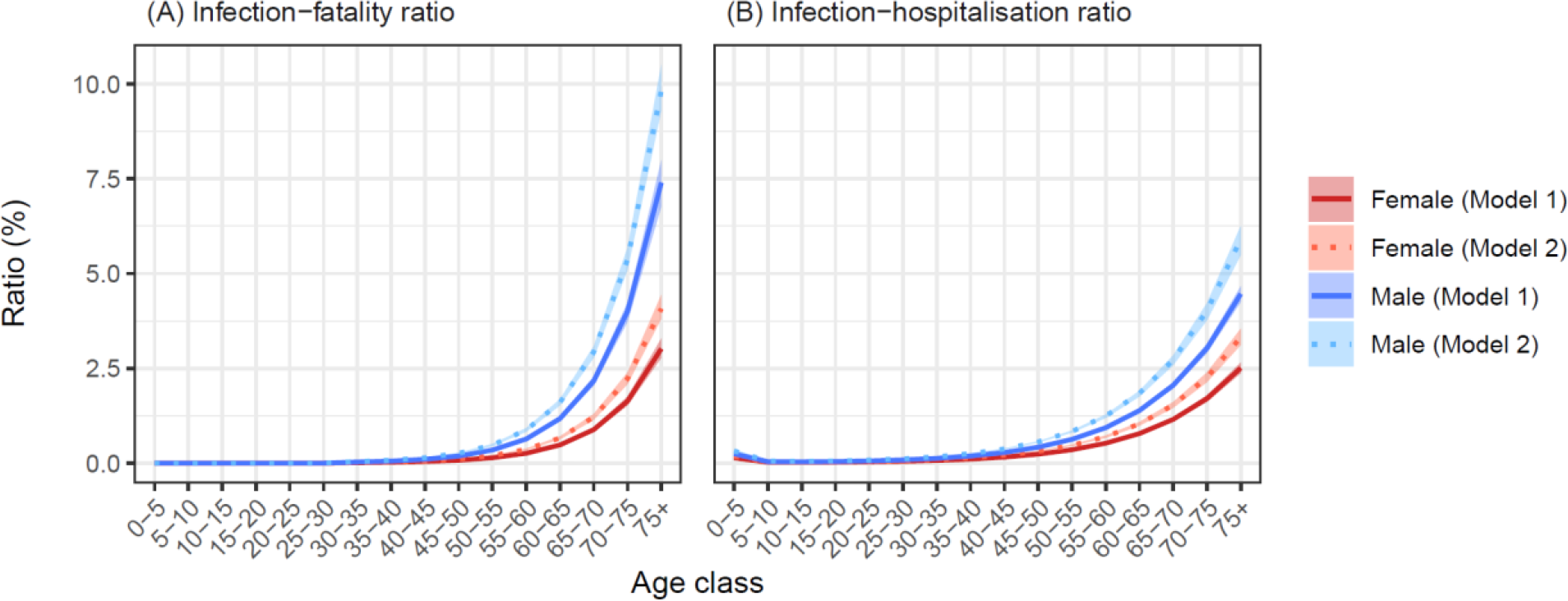
Estimated infection-fatality and infection-hospitalisation ratios. Model-based estimates (mean ± 95 % credible intervals) of age- and gender-structured (A) infection-fatality ratios (IFRs) and (B) infection-hospitalisation ratios (IHRs). Note that in panel (A), IFRs were fixed at zero for all age classes below 30 years.

Outcomes were similar for the second model which estimated *Q* = 24.5 [23.0, 25.1]% of the Manaus population was effectively removed from the susceptible pool. Relative to the baseline model, this second model estimated the expected offspring per case fell below one after April 11 when the attack rate was 46.8 [46.0, 47.7]%, and reduced the final attack rate to 65.0 [63.7, 65.6]%.

## Discussion

Early time-series modelling of the COVID-19 epidemic in Brazilian states up to 6 May 2020 used an unstructured model informed by state-level mobility indicators and reported deaths [12]. The model inferred population attack rates ranging from 0.13% to 10.6%, for Minas Gerais and Amazonas, respectively, and predicted ongoing epidemic growth throughout May. Other modelled *R*_*0*_ estimates based on reported COVID-19 cases from Amazonas state have similarly anticipated continued growth in cases through the month of June [13]. These models could neither explain nor accurately forecast observations of epidemic decline in Manaus.

Our analysis of excess deaths and hospitalisations in Manaus identifies rapid epidemic growth from late March, peaking in late April, with a return to baseline mortality in early June. These age- and gender-stratified models inferred heterogeneous transmission consistent with previous studies of SARS-CoV-1 [23] and SARS-CoV-2 [24]. In the baseline model, the expected offspring per case fell below one at a population-wide attack rate of 61.8%. This model suggests the herd-immunity threshold was exceeded in Manaus, with resulting attack rates inferred to be 86% by July 20 2020. In contrast, the attack rate stabilised at 65 % for the second model which estimated the risk of infection applied to *c*. 75% of the population only. Both models suggest that, primarily due to herd protection, the expected number of offspring per cases fell below one in early April.

Although both models estimated declining SARS-CoV-2 transmission over time due to herd immunity, their implications for the possibility of subsequent waves are qualitatively different. Although there are currently no data to support the assertion that some fraction of the Manaus population self-isolated, we consider the second model is likely more realistic because: (1) the estimated infection-fatality ratios are higher than reported from high-income countries [25] which we anticipate to be plausible; and (2) a population attack rate of 65% is more consistent with recent estimates based on a Manaus blood bank serosurvey [26] which reported the largest increase in seropositivity over the month of April.

We focus on Manaus as the epicentre of the COVID-19 epidemic in Amazonas, avoiding conflation of case numbers in this dense city of more than 2 million people with slower growing rural outbreaks across the state [6]. Given known information bias associated with limited early testing capacity, we used excess deaths and hospitalisations as the most objective indicators of epidemic activity presently available. Detailed cause-of-death data support the hypothesis that the majority of excess mortality over this time was COVID-19 related.

An important caveat is that estimated attack rates accumulate all infections and are agnostic to the presence or degree of symptoms. Severity of the clinical course of COVID-19 is associated with magnitude and persistence of the host immune response [27]. In consequence, our estimates of ‘exposure’ cannot be directly related to predicted antibody seroprevalence at the end of the first wave [28]. It is reasonably anticipated that population immunity will wane over time, requiring robust memory responses [29] to prevent reinfection or modify the clinical course [30].

Our findings support emerging evidence that population heterogeneity of SARS-CoV-2 transmission, attributable to a range of biological and sociological factors, reduces the herd immunity threshold [24, 31, 32]. Reduction of superspreading events through constraints on mixing group sizes is concordant with genomic studies showing successive extinction of imported strains under the influence of social and mobility restrictions [33].

While future infection clusters and outbreaks remain possible in Manaus due to unexposed subgroups, population-level immunity will likely constrain widespread transmission unless immunity wanes. As in other settings, underlying vulnerability of older age groups may be further exacerbated over time by reduced health seeking behaviours of individuals with pre-existing and new medical conditions [34]. Social measures should be informed by understanding of key enablers of superspreading and amplification, awareness of at-risk populations and tailored to context depending on population experience of the first wave.

Excess mortality data are not sufficiently timely to support real-time decision making, but in areas with limited testing may be a more reliable indicator of past epidemic activity than confirmed cases. To accurately support response to and assess the impact of the COVID-19 and other public health emergencies, improved access to diagnostics, and strengthening of reporting systems are needed in low- and middle-income settings. Cross-sectional and longitudinal seroprevalence studies are essential to understand markers and maintenance of immunity to inform prediction of long-term epidemiologic trends and bridging to likely vaccine impacts [26, 35].

## Data Availability

All data/code is provided as Supplementary Data with the paper. This will be available with the paper when published.

## Supplementary Material to

**Table S1.** Excess mortality in Manaus, Amazonas, during the COVID-19 epidemic (here defined as the period between March 19 and June 24, 2020).

| Age | Females |  |  | Males |  |  | Total |  |
| --- | --- | --- | --- | --- | --- | --- | --- | --- |
|  | Expected mortality | Observed mortality | Excess mortality | Expected mortality | Observed mortality | Excess mortality | Total Excess mortality | Excess mortality (%) |
| **0-5** | 84.5 | 65 | -19.5 | 109.6 | 83 | -26.6 | **-46.1** | **-23.7** |
| **5-10.** | 13.2 | 3 | -10.2 | 19.3 | 10 | -9.3 | **-19.5** | **-60** |
| **10-15.** | 13.5 | 6 | -7.5 | 19.3 | 11 | -8.3 | **-15.8** | **-48.2** |
| **15-20** | 26.1 | 14 | -12.1 | 64.3 | 45 | -19.3 | **-31.4** | **-34.7** |
| **20-25** | 24.1 | 22 | -2.1 | 116.4 | 78 | -38.4 | **-40.5** | **-28.8** |
| **25-30** | 26.7 | 30 | 3.3 | 92.2 | 81 | -11.2 | **-7.9** | **-6.6** |
| **30-35** | 30 | 35 | 5 | 81.2 | 88 | 6.8 | **11.8** | **10.6** |
| **35-40** | 38.3 | 53 | 14.7 | 79 | 118 | 39 | **53.7** | **45.8** |
| **40-45** | 44.6 | 88 | 43.5 | 72.4 | 154 | 81.7 | **125.1** | **107** |
| **45-50** | 53.3 | 110 | 56.7 | 79.8 | 184 | 104.2 | **160.9** | **120.8** |
| **50-55** | 59.2 | 114 | 54.8 | 99 | 225 | 126 | **180.8** | **114.3** |
| **55-60** | 73.2 | 180 | 106.8 | 121.2 | 310 | 188.8 | **295.6** | **152.1** |
| **60-65** | 95.6 | 215 | 119.4 | 139.2 | 436 | 296.8 | **416.2** | **177.3** |
| **65-70** | 101.6 | 283 | 181.4 | 148.6 | 510 | 361.4 | **542.8** | **216.9** |
| **70-75** | 110.2 | 276 | 165.8 | 143.6 | 456 | 312.4 | **478.2** | **188.4** |
| **75+** | 468.4 | 996 | 527.6 | 382.4 | 1208 | 825.6 | **1353.2** | **159.1** |
| **Total** | **1262.4** | **2490** | **1227.6** | **1767.5** | **3997** | **2229.5** | **3457.2** | **114.1** |

**Fig S1.**
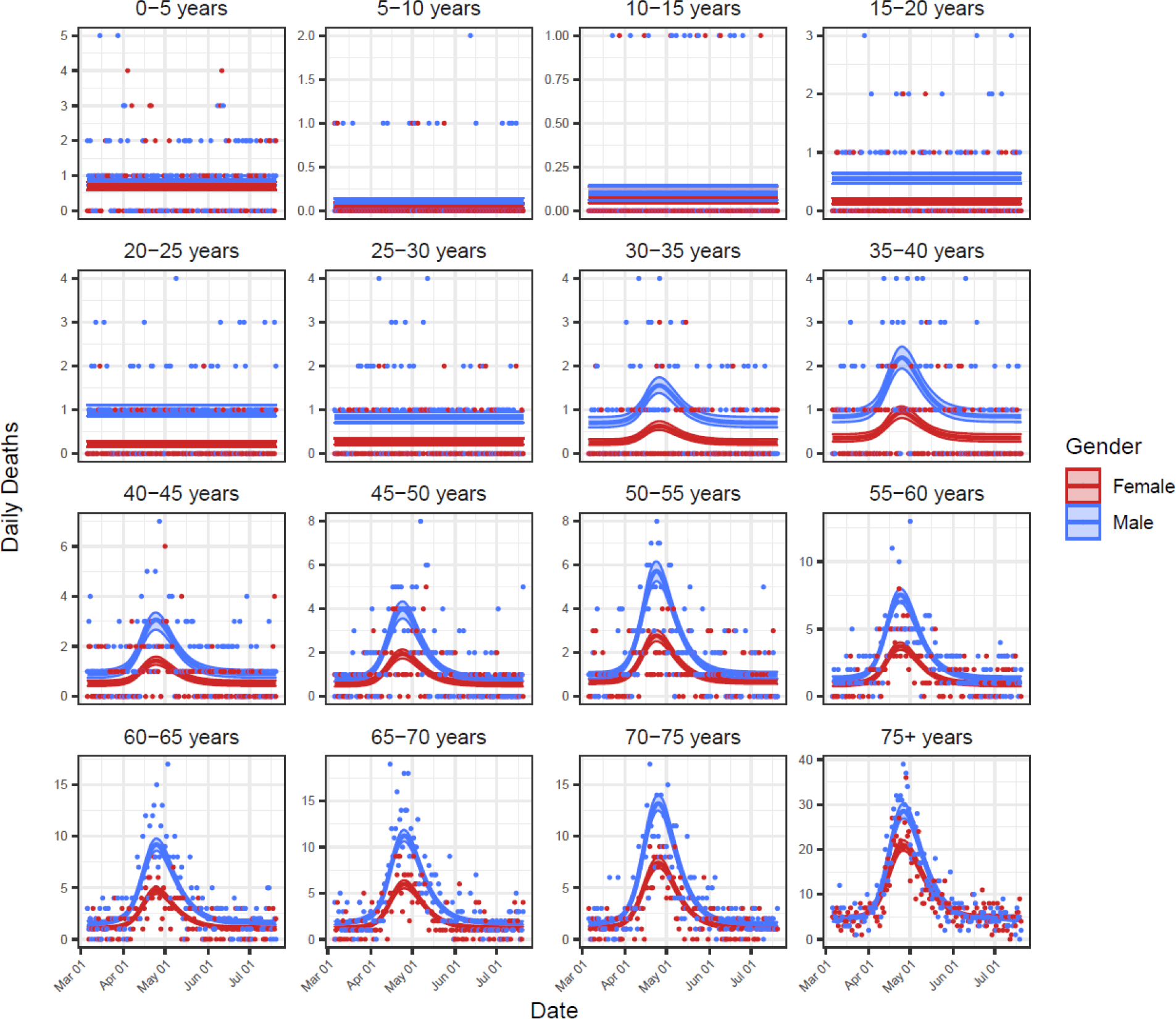
Baseline model fits to daily deaths data for all 32 age and gender classes. Shown are the model-inferred expected death rate from all causes (lines) ± 95% credible intervals (ribbons), and the mortality data used for model fitting (points).

**Fig S2.**
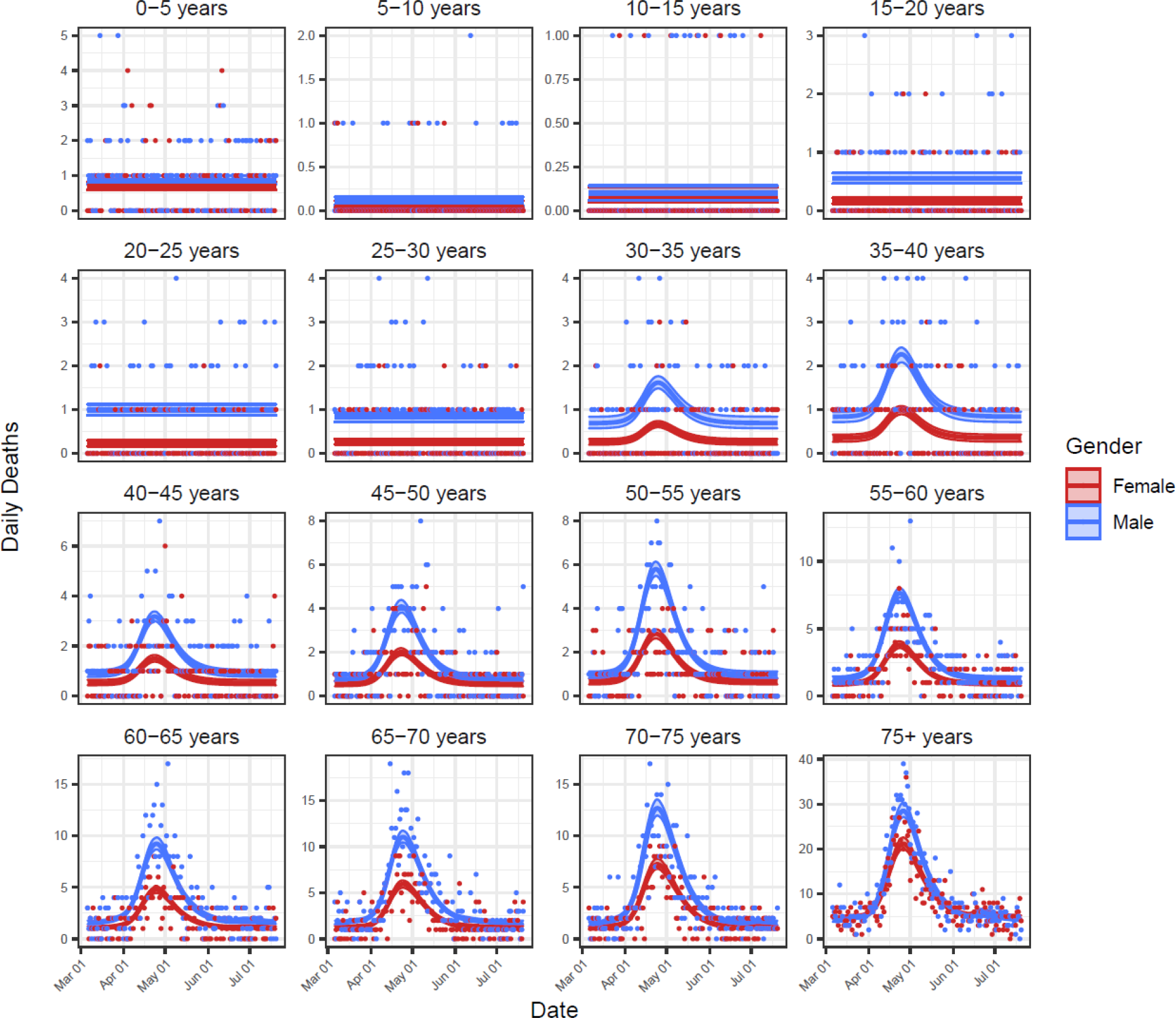
The fit of Model 2 to daily deaths data for all 32 age and gender classes. Shown are the model-inferred expected death rate from all causes (lines) ± 95% credible intervals (ribbons), and the mortality data used for model fitting (points).

**Fig S3.**
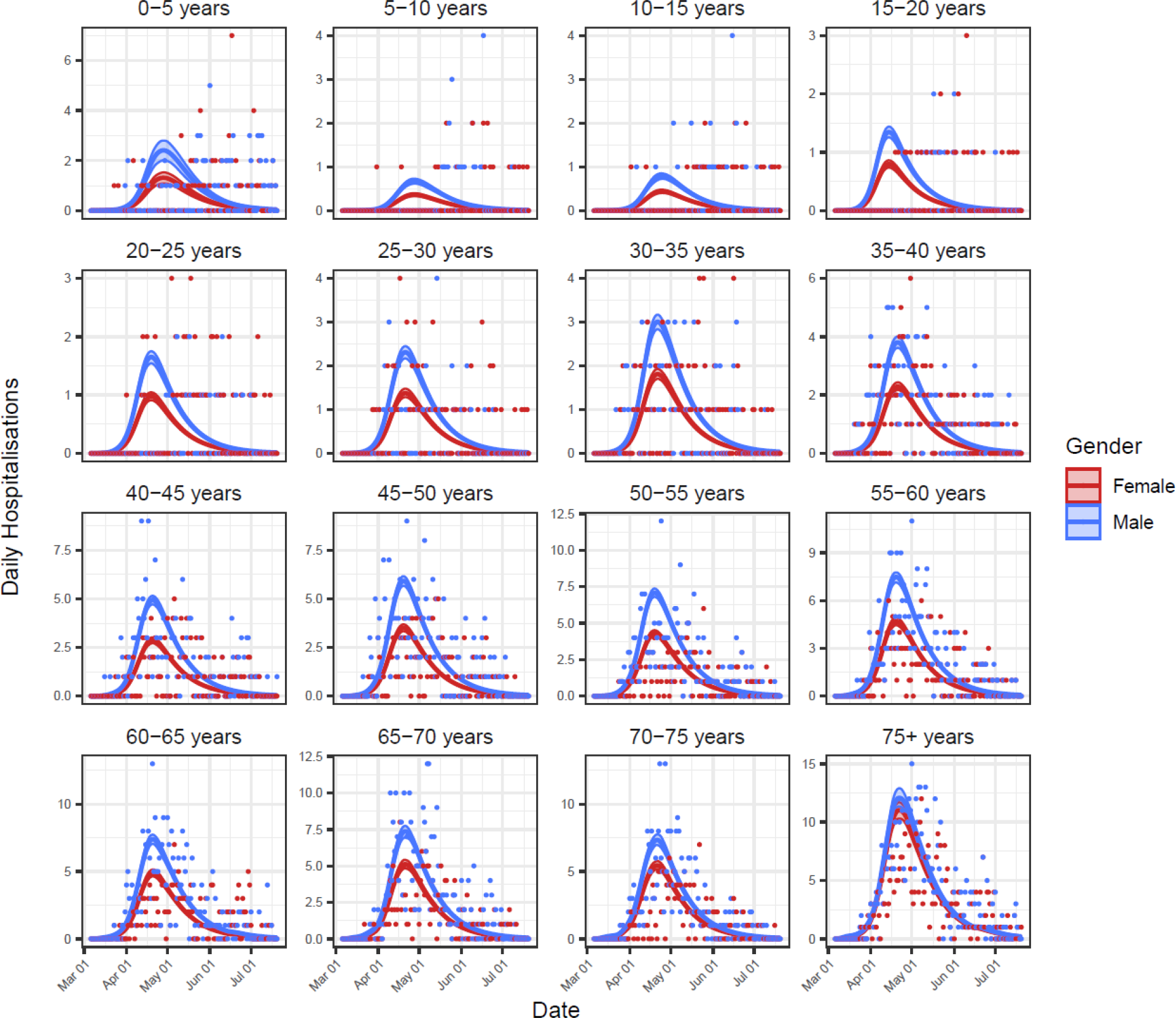
Baseline model fits to hospitalisations data for all 32 age and gender classes. Shown are the model-inferred expected hospitalisation rate (lines) ± 95% credible intervals (ribbons) and the hospitalisations data used for model fitting (points).

**Fig S4.**
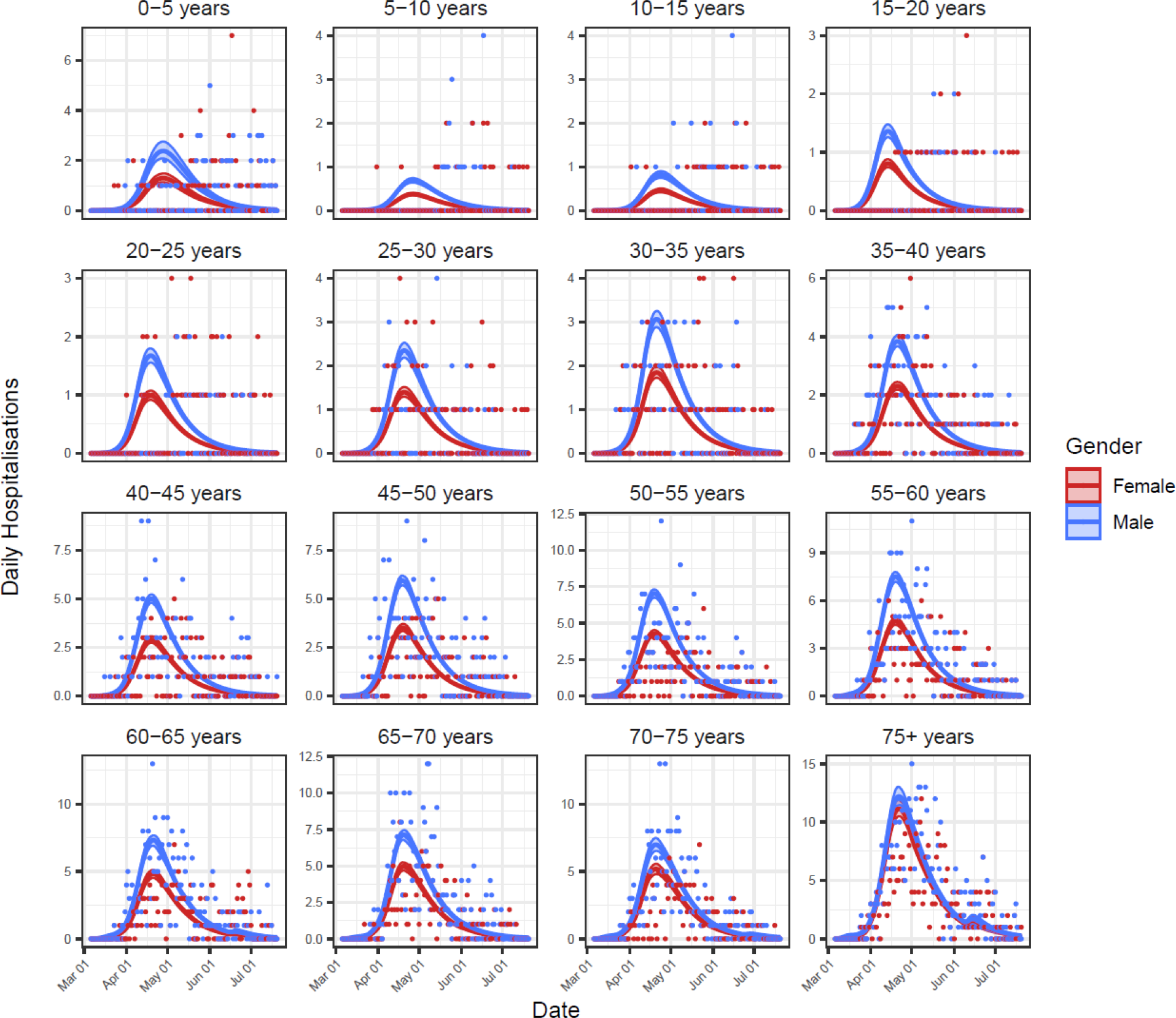
The fit of Model 2 to hospitalisations data for all 32 age and gender classes. Shown are the model-inferred expected hospitalisation rate (lines) ± 95% credible intervals (ribbons) and the hospitalisations data used for model fitting (points).

**Fig S5.**
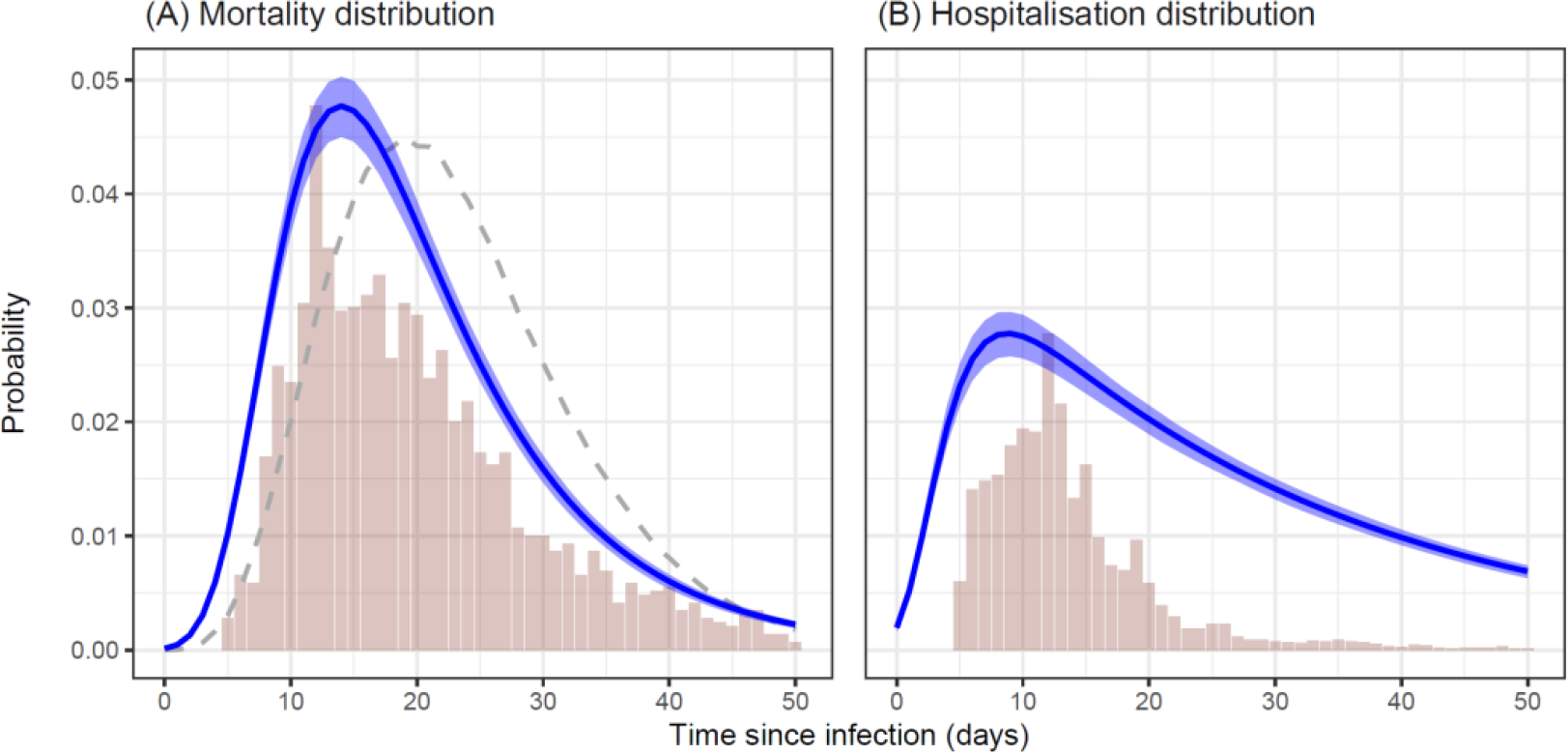
Estimates from the baseline model (mean ± 95 % credible intervals) of the shape of the (A) mortality distribution and (B) hospitalisation distribution, as a function of time since infection. Vertical bars indicate empirical frequencies derived from hospitalisation records for Manaus. In (A), the dashed line indicates the mortality distribution used by a previous COVID-19 modelling study for the State of Amazonas, Brazil [12].

**Fig S6.**
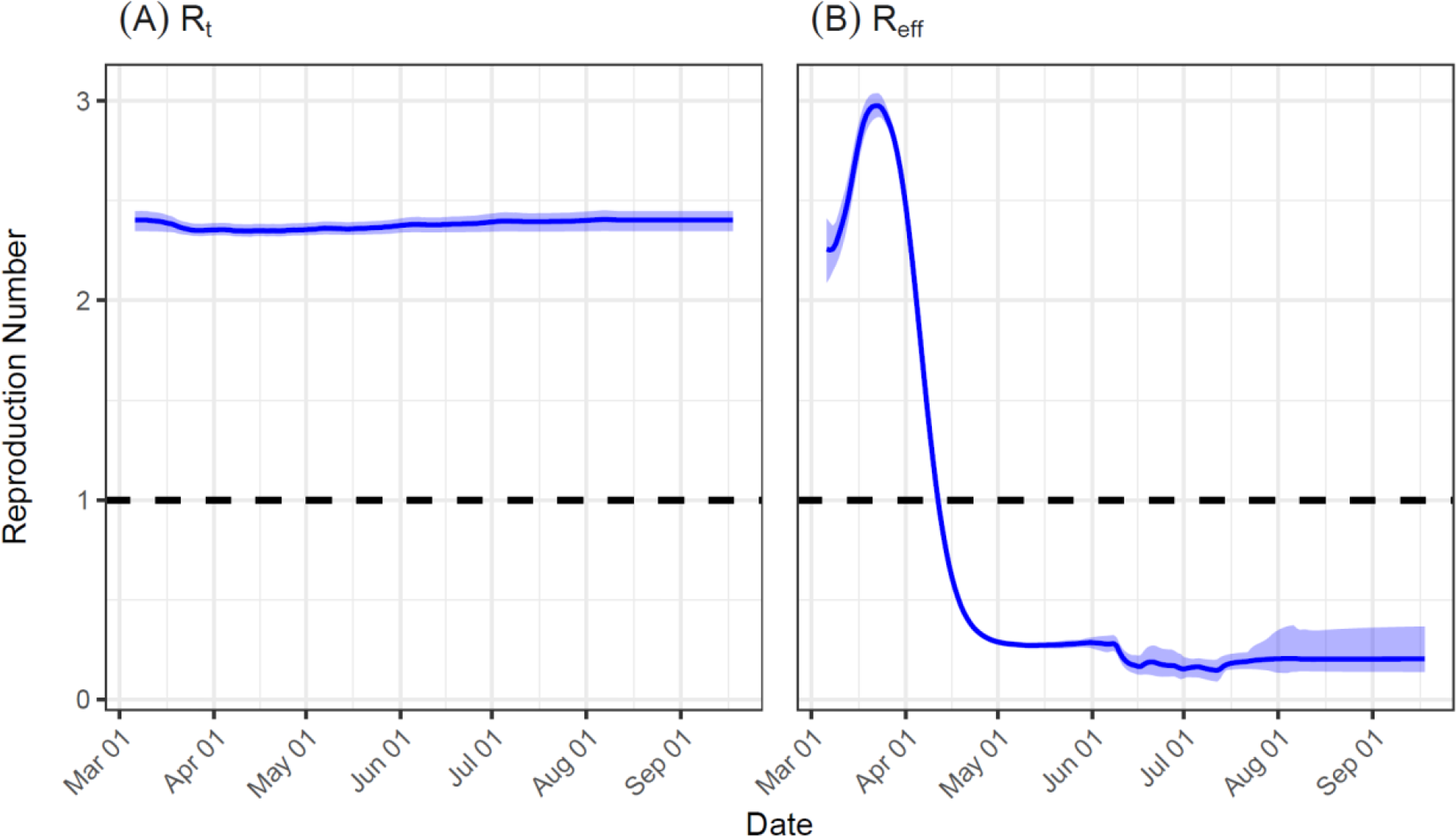
Estimates from the baseline model (mean ± 95 % credible intervals) of: (A) the impact of personal mobility on the time-varying reproduction number (*R*_*t*_); and (B) the population-level effective reproduction number (*R*_*eff*_) over time, which here is calculated as the sum of the products of all 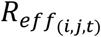 and the normalised case-infectivity vectors at each time *t*. In the latter, *R*_*eff*_ rises initially as the case distribution converges on the stable distribution, and *R*_*eff*_ < 1 indicates the switch to negative epidemic growth due to the development of herd immunity.

